# Combined assessment of *KRAS* mutational status and tumor size has no impact on prognosis in early-stage non-small cell lung cancer

**DOI:** 10.1101/2023.05.02.23289316

**Authors:** Ella A. Eklund, Ali Mourad, Clotilde Wiel, Sama I. Sayin, Henrik Fagman, Andreas Hallqvist, Volkan I. Sayin

## Abstract

**Background:** *KRAS* mutation status, stage and tumor size at the time of diagnosis are well-established independent prognostic factors in non-small cell lung cancer (NSCLC). Here, we investigate the prognostic value of combining survival data on *KRAS* mutation status and tumor size in early-stage NSCLC.

**Methods:** We studied the combined impact of *KRAS* mutational status and tumor size on overall survival (OS) and risk of death in patients with stage I-II NSCLC. We performed a retrospective study including 310 consecutively diagnosed patients with early (stage I-II) NSCLCs. All consecutive patients molecularly assessed and diagnosed between 2016-2018 with stage I-II NSCLC in the Västra Götaland region of western Sweden were included in this multi-center retrospective study. The primary study outcome was OS and risk of death (hazard ratio).

**Results:** Out of 310 patients with stage I-II NSCLC, 37% harbored an activating mutation in the *KRAS* gene. Our study confirmed staging and tumor size as prognostic factors. However, *KRAS* mutational status was not found to impact OS and there was no difference in the risk of death when combining *KRAS* mutational status and primary tumor size.

**Conclusions:** In our patient cohort, *KRAS* mutations in combination with primary tumor size are not associated with a worse prognosis in stage I-II NSCLC.

## Introduction

Non-small cell lung cancer (NSCLC) is the second most common cancer worldwide with 2.1 million new cases annually and the highest mortality rate with 1.8 million deaths. [1] Staging is a crucial aspect of NSCLC management, as it is one of the most important predictors of survival. The TNM staging system describes key tumor characteristics such as size, location, and whether the disease has spread to lymph nodes and/or distant organs [2-5]. There are four main stages in NSCLC (stage I-IV), with stage IV having the worst prognosis. Pathological stage is considered the most important prognostic factor for resected patients, with 5-year survival rates, gradually decreasing across stages, of 83% for stage IA, 71% for IB, 57% for IIA, 49% for IIB, 36% for IIIA, and 23% for IIIB [4].

The most frequent oncogenic driver in NSCLC is the Kirsten rat sarcoma viral oncogene (*KRAS*), which is present in up to 40% of all cases, with the most common mutations being *G12C, G12V*, and *G12D* [6]. *KRAS* mutations are associated with worse outcomes after chemotherapy and radiotherapy, with shorter OS in stage III and IV patients [7-14]. In early-stage NSCLC, however, while several studies have shown that *KRAS* mutations negatively influence the prognosis [15-17], others have shown no significant effect [18]. Most recently, it was reported that *KRAS G12C* mutation (but not other *KRAS* mutations or with no mutation in *KRAS*) significantly increased risk of disease recurrence in stage I surgically resected lung adenocarcinomas [19]. However, while the study found this in two distinct local cohorts of IRE-LUAD (Rome, Italy) and MSK-LUAD (New York, USA), data extracted from The Cancer Genome Atlas (TCGA) showed no significant difference. Hence, the debate about the prognostic value of *KRAS* mutational status in early NSCLC is ongoing [20, 21]. In fact, given the lack of consensus regarding its effects on prognosis, testing for *KRAS* mutations for resectable stage I and II tumors is currently not recommended in clinical guidelines [22]. In addition, several inhibitors that specifically bind *KRAS-G12C* have been investigated in clinical trials, with sotorasib becoming the first treatment to gain approval for adults with stage IV NSCLC harboring a *KRAS-G12C* mutation as second-line therapy [23-27]. However, treatment with sotorasib is not currently recommended for patients with early-stage NSCLC due to lack of evidence showing positive outcomes of treatment in this group.

Therefore, further investigations are warranted to identify potential subgroups in Stage I-III disease who may still have to gain from effective and well-established treatments, and to add to the pool of clinical data required to study this further. One strategy is to stratify patients according to *KRAS* mutational status together with other key prognostic factors, such as tumor size. Primary tumor size is an established prognostic factor in NSCLC, with larger tumors being associated with poorer survival [21, 28-31]. The reason for this association is not yet fully understood but larger tumors may be more resistant to therapy due to having poorer blood supply, differential metabolism, and potentially a higher likelihood of micrometastatic disease compared to smaller tumors [32-35]. Further research is needed to elucidate the underlying mechanisms. However, when considering primary tumor size, the grouping as early (I-II), advanced (III), and metastasized (IV) NSCLC can be argued to be more clinically relevant due to that stage I-II is primarily based on tumor size whereas a spread to the lymph nodes, a negative prognostic factor, is more common in stage III [3, 21].

To our knowledge, no one has investigated the combined impact of primary tumor size and *KRAS* mutational status on OS and risk of death in stage I-II NSCLC. However, in Sweden, reflex testing for targetable alterations in NSCLC, including *KRAS* mutational status, has been widely implemented since 2015 for all stages. By including all consecutive patients diagnosed with stage I-II NSCLC and molecularly assessed between 2016-2018 in Västra Götaland, the second largest county in Sweden with a population of 1.7 million, the current retrospective cohort study provides a unique real-world dataset for assessing the impact of combining *KRAS* mutations with primary tumor size.

To summarize, primary tumor size is a key determinant of prognosis especially in the early stages of NSCLC. At the same time, the prognostic value of *KRAS* mutational status in early disease stages remains unclear. Hence patients diagnosed at an early stage are not automatically tested for *KRAS* mutations and recommended treatment with *KRAS*-targeted therapy. Here, we investigate whether there is prognostic value in combining *KRAS* mutational status with tumor size to aid in clinical stratification of potentially treatment-responsive subgroups in early-stage NSCLC.

## Materials and Methods

### Patient population

We conducted a multi-center retrospective study including all consecutive NSCLC patients diagnosed with stage I-II NSCLC and molecular assessment performed between 2016-2018 in Västra Götaland, Sweden (*n* = 310). Further inclusion criteria included the availability of tumor size from CT scanning or a pathology report as well as follow-up data. Patients were excluded if diagnosed before 2016, had no digitally accessible patient charts, no tumor measurements noted in the patient charts, or had recurrent disease.

Patient demographics (age, sex, Eastern Cooperative Oncology Group [ECOG] performance status [PS], and smoking history), cancer stage, pathological details (histology, mutational status including *KRAS* mutational status and subtype), first-line treatment and outcome data were retrospectively collected from patient charts and the Swedish Lung Cancer Registry. Clinical staging was based on TNM staging guidelines 7^th^ edition [4]. TNM staging 8^th^ edition released in 2017 was introduced in Swedish guidelines in 2018, and full implementation was reached in 2019. Ethical approval was obtained from the Swedish Ethical Review Authority prior to study commencement (Dnr 2019-04771 and 2021-04987). No informed consent was required due to all data presented in a de-identified form according to the Swedish Ethical Review Authority.

### Mutational status

Patients were assessed with next-generation sequencing (NGS) for mutational status on DNA from formalin-fixed paraffin-embedded (FFPE) blocks or cytological smears using the Ion AmpliSeq™ Colon and Lung Cancer Panel v2 from Thermo Fisher Scientific as part of the diagnostic workup process at the Department of Clinical Pathology at Sahlgrenska University Hospital, assessing hotspot mutations in *EGFR, BRAF, KRAS*, and *NRAS*. Until June 2017, *ALK*-fusions were assessed with immunohistochemistry (IHC), and with fluorescence in situ hybridization (FISH) if positive or inconclusive IHC. *ROS1* was analyzed upon request with FISH. Thereafter, *ALK, ROS1*, and *RET* fusions were assessed on RNA using the Oncomine Solid Tumor Fusion Panel from Thermo Fisher Scientific.

### Tumor size

To obtain the most recent and accurate untreated primary tumor size, measurements were collected from the radiology report of computed tomography (CT) performed before a final diagnosis of NSCLC was established; this is referred as clinical staging. In patients who underwent surgical resection, the actual primary tumor size was also collected from the pathology report, also referred as pathological staging (PAD). The largest tumor diameter was collected and reported in millimeters.

### Study objectives

The primary outcome of this study was OS and risk of death, defined as the interval between the date of first treatment and the date of death from any cause. Patients alive or lost to follow-up at the cut-off date were censored at last contact. Median follow-up time was estimated using the reverse Kaplan-Meier method. We compared OS and risk of death stratified by *KRAS* mutational status, i.e., with no mutation in *KRAS* (wildtype, *KRAS*^WT^), all *KRAS* mutations (*KRAS*^MUT^), *KRAS G12C* mutations (*KRAS*^MUT G12C^) and all *KRAS* mutations other than *G12C* (*KRAS*^MUT not G12C^).

### Statistical analysis

Clinical characteristics were summarized using descriptive statistics and evaluated with univariate analysis in table form. Survival was estimated using the Kaplan-Meier method. The log-rank test was used to assess significant differences in OS between *KRAS*^WT^ and *KRAS*^MUT^ groups. To evaluate if there was a significant difference in primary tumor size between *KRAS*^MUT^ and *KRAS*^WT^, the Mann-Whitney U test was used. Cox proportional hazard regression was conducted to measure the influence of tumor size on the risk of death (hazard ratio [HR]) stratified by *KRAS* mutational status. We defined an interaction term between tumor size (largest diameter in mm) and *KRAS* mutational status to assess the combined impact on the risk of death (HR). Statistical significance was set at *p* < 0.05 and no adjustments were made for multiple comparisons. Data analysis was conducted using IBM SPSS Statistics version 27 and GraphPad Prism version 9.

## Results

### Patients and tumor characteristics

A total of 310 consecutive patients, who were diagnosed with stage I-II NSCLC during 2016-2018 in Västra Götaland, Sweden and for whom genetic data was available, were included in this retrospective cohort study (Fig. 1). In the total population, majority of patients were female (187, 60.3%), with a median age of 70 years, and most were current or former smokers (267, 86%) (Table 1). Most patients had good PS with ECOG 0-1 at diagnosis (285, 92%) and the proportion of N1 was low (18, 5.8%). NSCLC was predominantly adenocarcinoma of the lung (281, 90.6%), while squamous cell carcinoma incidence was relatively low (11, 3.5%), which was expected due to the selection of histological type for NGS assessment. Of included patients, over a third (115, 37%) had a *KRAS* mutation (Table 1). This percentage matches what has been previously reported [9], showing good representativeness of the patient group studied here. When comparing the baseline characteristics of *KRAS*^WT^ with *KRAS*^MUT^ patients, a greater proportion of those with *KRAS*^MUT^ were female and current or former smokers. There were no cases of squamous cell carcinoma in the *KRAS*^MUT^ group. The most common KRAS mutation was G12C (47%). In the total population, majority of patients underwent surgical resection (273, 88%; Table 2). Three patients did not receive any treatment and were excluded from further survival analyses. Median follow-up time was 63 months (95% CI, 59.7-68.3) and the data cut-off date was 31 October 2022.

**Table 1.** Characteristics of the total cohort as well as stratified by *KRAS*^WT^ and *KRAS*^MUT^. Data are presented as n (%). ECOG PS, Eastern Cooperative Oncology Group Performance Status. T, Tumor. N, Nodulus.

|  | **Total** | **KRAS WT** | **KRAS MUT** |
| --- | --- | --- | --- |
|  | n (%) | n (%) | n (%) |
| Total | 310 (100) | 195 (63.0) | 115 (37.0) |
| Age in years, median (range) | 70 (35-85) | 70 (35-85) | 70 (48-84) |
| *Sex* |  |  |  |
| Male | 123 (39.7) | 88 (45.1) | 35 (30.4) |
| Female | 187 (60.3) | 107 (54.9) | 80 (69.6) |
| *Smoking history* |  |  |  |
| Current smoker | 99 (31.9) | 51 (26.2) | 48 (41.7) |
| Former smoker | 168 (54.2) | 106 (54.4) | 62 (53.9) |
| Never smoker | 43 (13.9) | 38 (19.5) | 5 (4.3) |
| *Performance status* |  |  |  |
| ECOG 0 | 144 (46.5) | 82 (42.1) | 62 (53.9) |
| ECOG 1 | 141 (45.5) | 96 (49.2) | 45 (39.1) |
| ECOG 2 | 24 (7.7) | 16 (8.2) | 8 (7.0) |
| ECOG 3 | 1 (0.3) | 1 (0.5) | 0 |
| ECOG 4 | 0 | 0 | 0 |
| *Histology* |  |  |  |
| Adenocarcinoma | 281 (90.6) | 168 (86.2) | 113 (98.3) |
| Squamous cell carcinoma | 11 (3.5) | 11 (5.6) | 0 |
| NCSLC NOS | 18 (5.9) | 16 (8.2) | 2 (1.7) |
| *Mutation status* |  |  |  |
| None known | 124 (40.2) | 124 (63.6) |  |
| KRAS | 115 (37.0) | 0 |  |
| EGFR | 54 (17.4) | 54 (27.7) |  |
| BRAF | 6 (1.9) | 6 (3.1) |  |
| ALK | 4 (1.3) | 4 (2.1) |  |
| ROS1 | 3 (1.0) | 3 (1.5) |  |
| RET | 1 (0.3) | 1 (0.5) |  |
| Other | 3 (1.0) | 3 (1.5) |  |
| *KRAS submutation* |  |  |  |
| G12A |  |  | 9 (7.8) |
| G12C |  |  | 54 (47.0) |
| G12D |  |  | 15 (13.0) |
| G12V |  |  | 26 (22.6) |
| Other |  |  | 11 (9.6) |
| *TNM* |  |  |  |
| T1a | 99 (31.9) | 55 (28.2) | 44 (38.3) |
| T1b | 76 (24.5) | 54 (27.7) | 22 (19.1) |
| T1c | 12 (3.9) | 7 (3.6) | 5 (4.3) |
| T2a | 67 (21.6) | 46 (23.6) | 21 (18.3) |
| T2b | 28 (9.0) | 15 (7.8) | 13 (11.3) |
| T3 | 28 (9.0) | 18 (9.2) | 10 (8.7) |
| N0 | 292 (94.2) | 185 (94.9) | 107 (93.0) |
| N1 | 18 (5.8) | 10 (5.1) | 8 (7.0) |
| *Measurement modality* |  |  |  |
| CT-scan (mm) | 305 (98.4) | 190 (97.4) | 115 (100) |
| PAD | 273 (88.0) | 171 (88.7) | 102 (88.7) |
| *At last follow up 31/10-2022* |  |  |  |
| Alive | 206 (66.5) | 128 (65.6) | 78 (67.8) |
| Deceased | 104 (33.5) | 67 (34.4) | 37 (32.2) |
| *Survival* |  |  |  |
| *Mean survival (months)* | 63 | 62 | 64 |

**Table 2.** Summary of first-line treatments in the total cohort as well as stratified by *KRAS*^WT^ and *KRAS*^MUT^. Data are presented as n (%).

|  | **Total** | **KRAS WT** | **KRAS MUT** |
| --- | --- | --- | --- |
|  | n (%) | n (%) | n (%) |
| Total | 310 (100) | 195 (63.0) | 115 (37.0) |
| Surgery | 273 (88.0) | 171 (87.7) | 102 (88.7) |
| Curative chemoradiotherapy | 7 (2.3) | 6 (3.1) | 1 (0.9) |
| Medical treatment | 2 (0.6) | 1 (0.5) | 1 (0.9) |
| Stereotactic radiotherapy | 11 (3.5) | 11 (5.6) | 0 (0) |
| Radiotherapy | 14 (4.5) | 3 (1.5) | 11 (9.6) |
| No treatment | 3 (1.0) | 3 (1.5) | 0 (0) |

**Figure 1.**
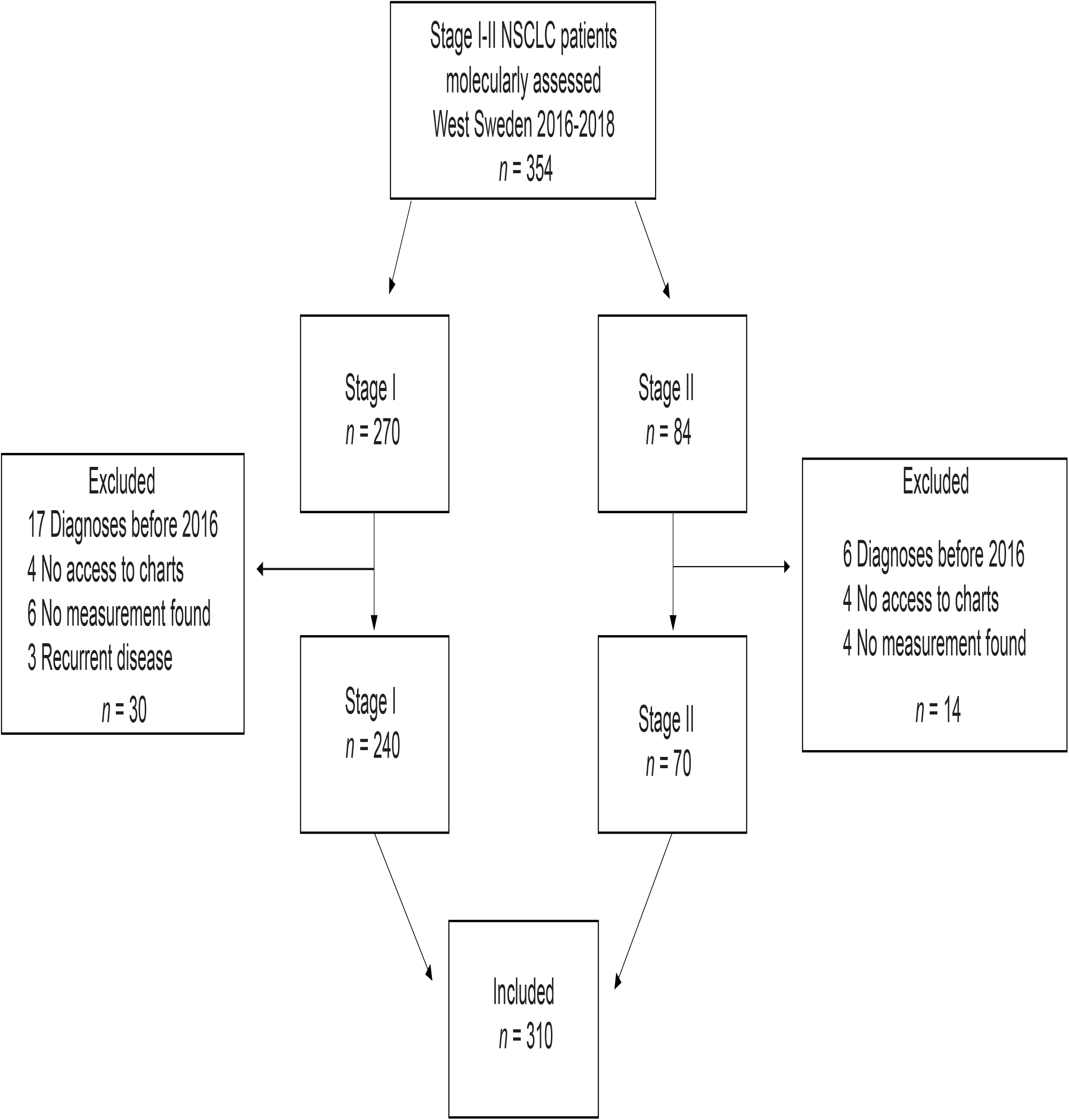
Patient selection. Flow chart showing patient selection for the study.

### No significant difference in survival for all patients stratified by *KRAS* mutations

When comparing OS for all (stage I-II) patients stratified by *KRAS* mutational status, no significant difference was detected with a mean OS (median not reached) of 74 months for *KRAS*^WT^ vs 63 months for *KRAS*^MUT^ (*p* = 0.847; Fig 2A). Further stratification of the *KRAS* mutated group by the G12C mutation also did not significantly change survival: 74 months for *KRAS*^WT^, 61 months for *KRAS*^MUT not G12C^ and 63 months for *KRAS*^MUT G12C^ (*p* = 0.834; Fig 2D).

**Figure 2.**
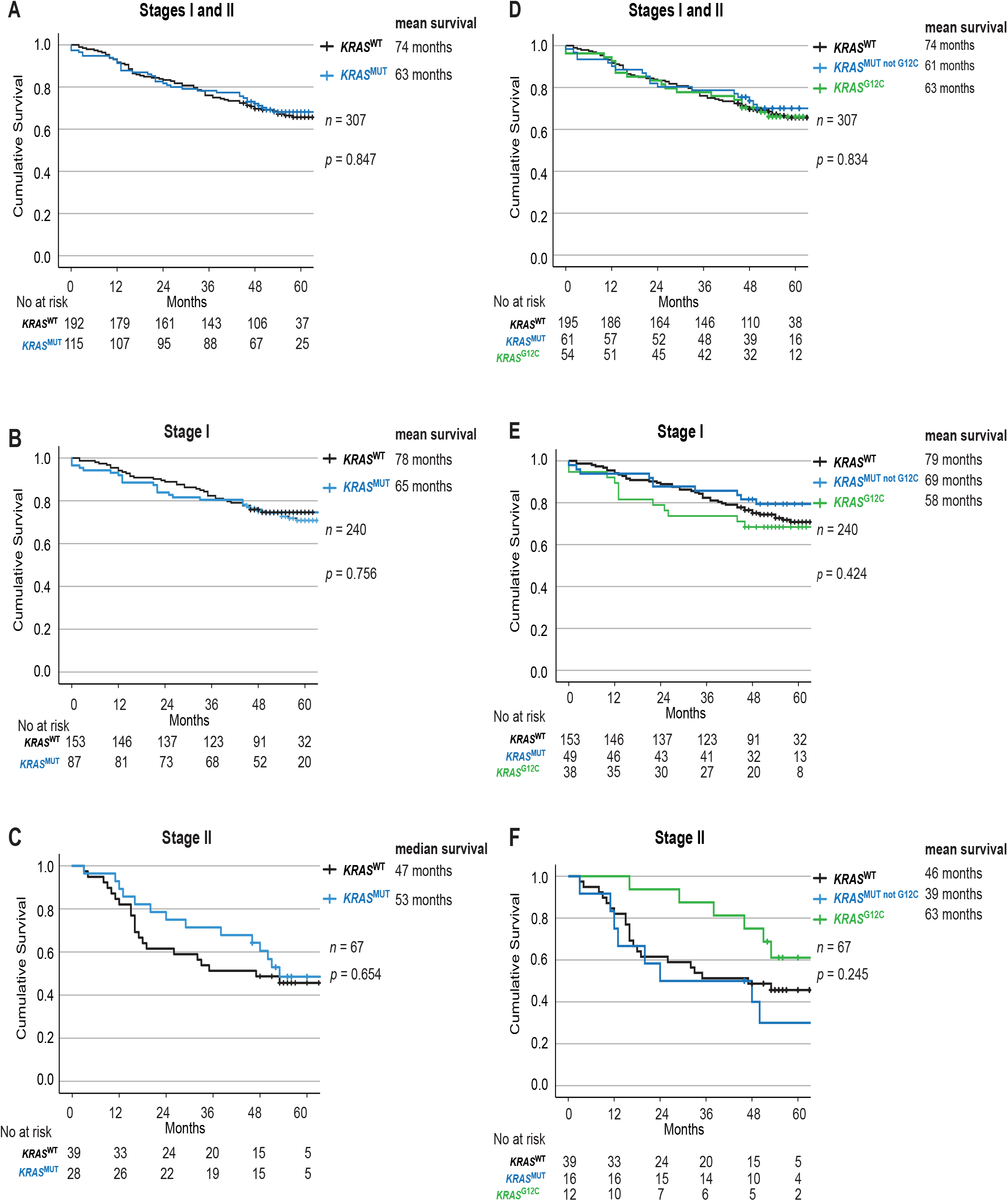
Impact of *KRAS* mutational status on overall survival in Stage I and II NSCLC. Kaplan-Meier estimates comparing overall survival between (**A, D**) all patients, (**B, E**) Stage I and (**C, F**) Stage II patients with no mutation in *KRAS* (wildtype, *KRAS*^WT^), with all *KRAS* mutations (*KRAS*^MUT^), only *KRAS*-*G12C* mutations (*KRAS*^MUT G12C^) and *KRAS* mutations other than *G12C* (*KRAS*^MUT not G12C^).

### No significant difference in survival for patients in Stage I or Stage II disease combined with *KRAS* mutations

There were also no significant differences according to *KRAS* mutational status in stage I (Fig 2B, 2E) or Stage II (Fig 2C, 2F). Similarly in resected patients, no significant difference was observed with a mean OS (median not reached) of 78 months for *KRAS*^WT^ vs 65 months for *KRAS*^MUT^ (*p* = 0.856; Supplemental Fig. 1A), or between the subgroups of *KRAS*^MUT^ (*p* = 0.471; Supplemental Fig.1B).

Next, we stratified by stage and found mean OS (median not reached) of 79 months for stage I vs 50 months for stage II (Fig. S2A). We then conducted the analysis separately according to *KRAS* mutational status. For *KRAS*^WT^, the mean OS (median not reached) was 78 months for stage I vs 46 months for stage II (Fig. S2B), and for *KRAS*^MUT^, 65 months for stage I vs 53 months for stage II (Fig. S2C).

### No significant difference in survival for patients with TNM-stage T1, T2 or T3 disease combined with *KRAS* mutations

Next, we stratified patients using T-staging and studied OS according to *KRAS* mutational status. Among those with T1 disease, *KRAS*^WT^ had 83 months while *KRAS*^MUT^ had 66 months OS (*p* = 0.751; Fig 3A). Further, *KRAS*^MUT not G12C^ patients had survival of 70 months and *KRAS*^MUT G12C^ had 61 months (*p* = 0.344; Fig 3D). In the T2 group, *KRAS*^WT^ had 53 months while *KRAS*^MUT^ had 59 months OS (*p* = 0.495; Fig 3B). *KRAS*^MUT not G12C^ patients had survival of 51 months and *KRAS*^MUT G12C^ had 66 months (*p* = 0.389; Fig 3E). Similarly, in the T3 group, *KRAS*^WT^ had 47 months while *KRAS*^MUT^ had 50 months OS (*p* = 0.966; Fig 3C). *KRAS*^MUT not G12C^ patients had survival of 50 months and *KRAS*^MUT G12C^ had 53 months (*p* = 0.984; Fig 3F).

**Figure 3.**
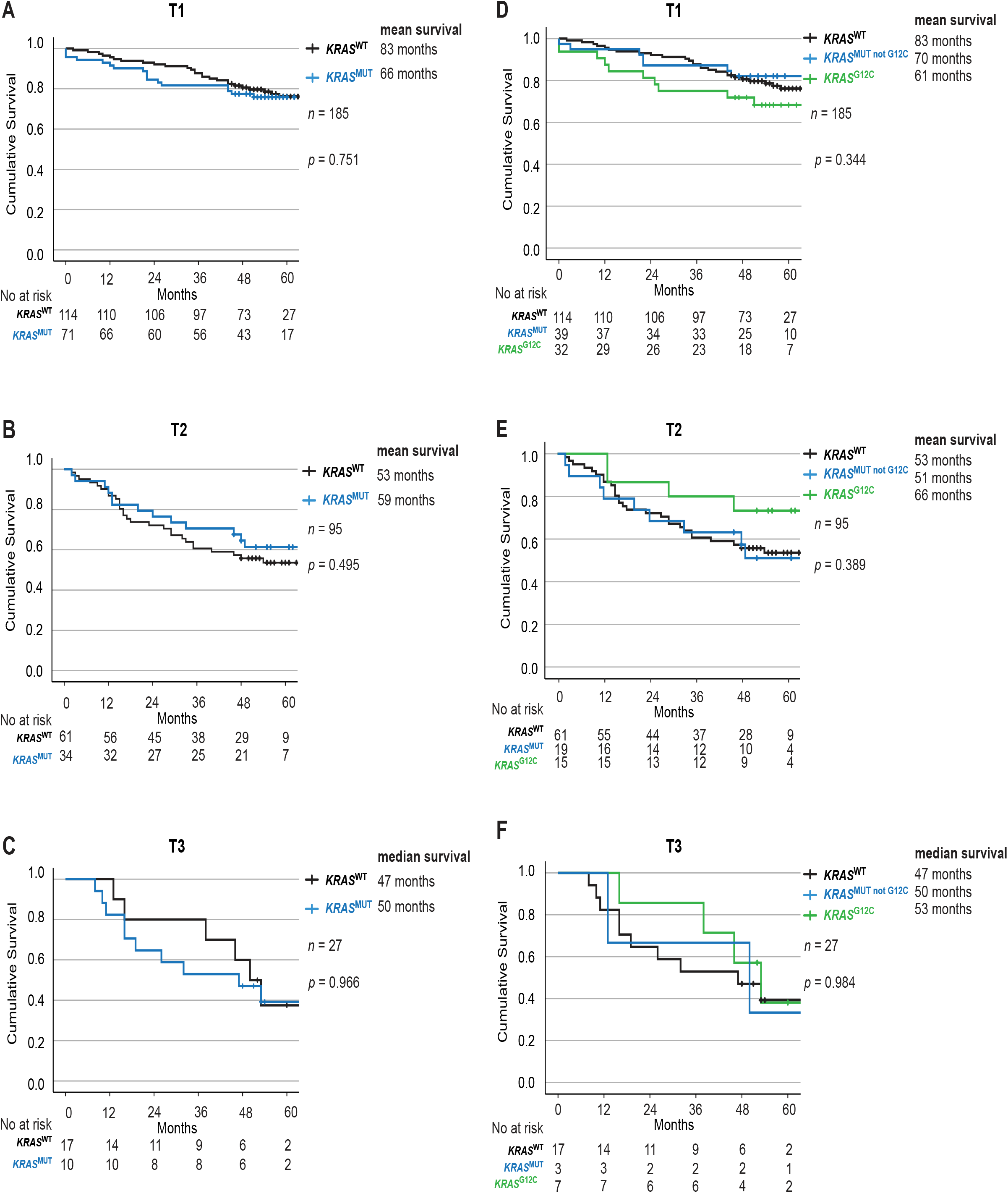
Impact of *KRAS* mutational status on overall survival across TNM-stages in NSCLC. Kaplan-Meier estimates comparing overall survival between (**A, D**) T1, (**B, E**) T2 and (**C, F**) T3 patients with no mutation in *KRAS* (wildtype, *KRAS*^WT^), with all *KRAS* mutations (*KRAS*^MUT^), only *KRAS*-*G12C* mutations (*KRAS*^MUT G12C^) and *KRAS* mutations other than *G12C* (*KRAS*^MUT not G12C^).

We further analyzed the impact of T stage on survival and found that it correlated as expected with mean OS of 82 months for T1, 55 months for T2, and 46 months for T3 (*p* < 0.001; Fig. S3A). The same trend was observed when separately analyzing *KRAS*^WT^ with a mean OS (median not reached) of 83 months for T1, 53 months for T2, and 45 months for T2 (*p* < 0.001; Fig. S3B), and *KRAS*^MUT^ with a mean OS of 65 months for T1, 58 months for T2, and 48 months for T3 (*p* < 0.023; Fig. S3C).

### *KRAS* mutations are associated with smaller tumor size measured from CT scans, but not resection specimens/PAD

To evaluate differences between primary tumor size from CT scans at diagnosis stratified by *KRAS* mutational status, we used the Mann-Whitney U test. The test revealed that *KRAS*^MUT^ primary tumors were significantly smaller at diagnosis, with a median size of 20 mm (*n* = 115) vs *KRAS*^WT^ primary tumors with a median size of 25 mm (*n* = 190) (*p* = 0.043; Fig. 4A). However, when looking at tumor size as assessed in resected specimens, there were no differences; *KRAS*^WT^ median size 22 mm (*n* = 171) vs *KRAS*^MUT^ median size 21 mm (*n* = 102) (*p* = 0.16; Fig. 4B).

**Figure 4.**
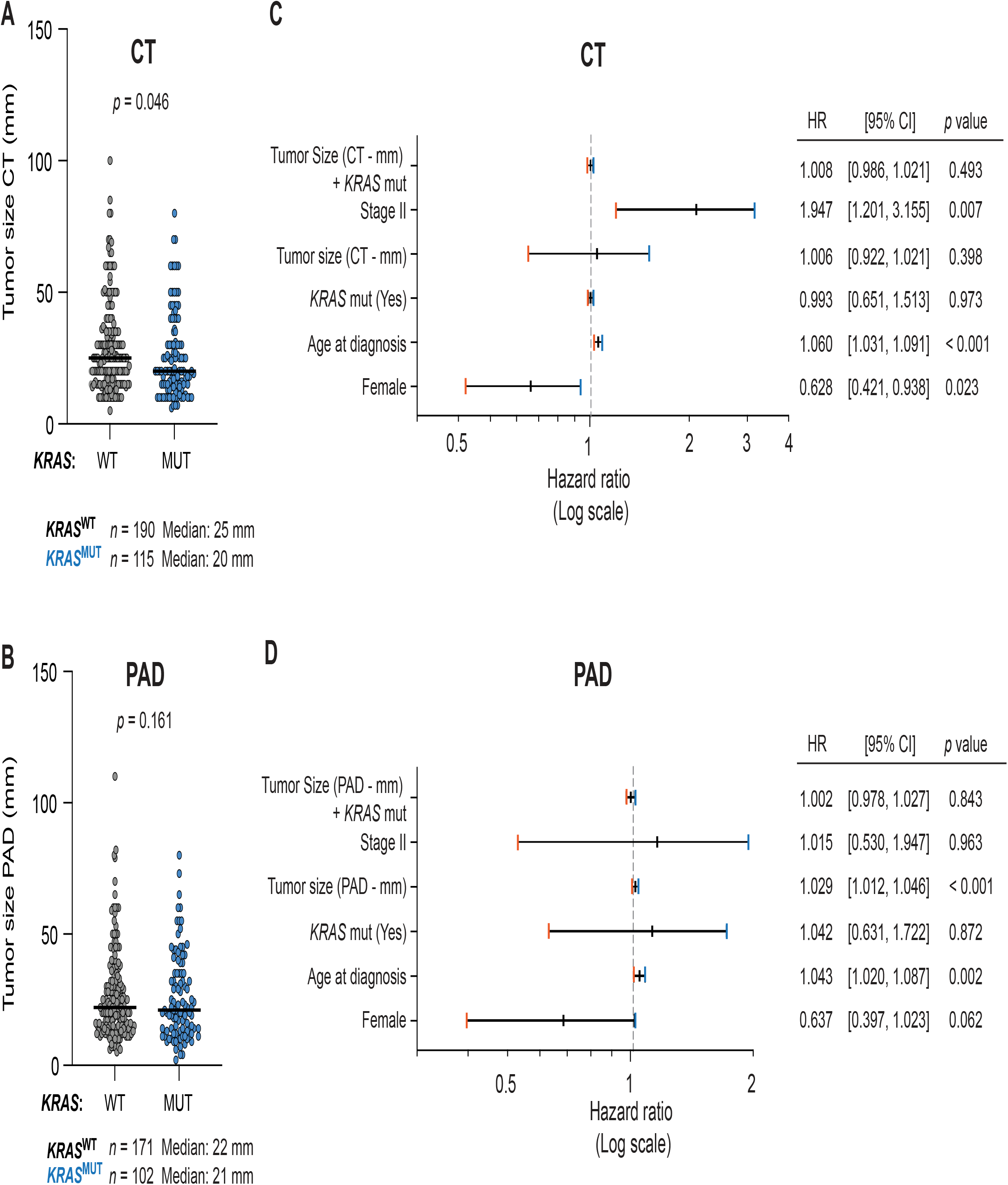
KRAS mutations are associated with smaller tumor size at diagnosis. Primary tumor size from (**A**) CT scans and (**B**) resection specimens (PAD) in patients with no mutation in *KRAS* (wildtype, *KRAS*^WT^) or with *KRAS* mutations (*KRAS*^MUT^). Forest plot of multivariate COX regression analysis for patients with tumor size from (**C**) CT scan and (**D**) resection specimens (PAD).

### Larger tumor size measured from resection specimens/PAD, but not CT scans, is associated with a higher risk of death

We found that increase in primary tumor size determined from CT scans did not have a significant effect on risk of death (HR, 1.006; 95% CI, 0.922-1.021; *p* > 0.5) (Fig 4C). However, when testing the correlation between primary tumor size as assessed in resection specimens, we found a significantly increased risk of death (HR, 1.029; 95% CI, 1.012-1.046; *p* < 0.001) (Fig 4D). The risk of death increases with 2.9% for every mm increase of size.

### The combination of *KRAS* mutational status and tumor size does not impact the risk of death

To test if the combination of tumor size and *KRAS* mutational status impacts the risk of death, we defined an interaction term including both variables. For primary tumor size from CT scans and *KRAS* mutational status, no significant difference in the risk of death was detected (HR, 1.008; 95% CI, 0.988-1.030; *p* > 0.5) (Fig 4C). Similarly, there were no significant differences for primary tumor size and *KRAS* mutational status when measured in resection specimens (HR, 1.002; 95% CI, 0.978-1.027; *p* = 0.807) (Fig 4D).

## Discussion

In this study, we assessed the prognostic value of combining *KRAS* mutational status with tumor size in early-stage NSCLC. We found that combining these variables had no significant effect on overall survival or the risk of death.

In alignment with previous findings, we found in our patient cohort that later disease stage and larger primary tumor size is associated with worse survival. Interestingly, we found that these correlations are sustained independent of *KRAS* mutational status. Importantly, the established literature on how KRAS mutations affect outcomes in early-stage NSCLC is varying between worse survival and no significant difference. Here, we find as the latter, that *KRAS* mutational status alone does not significantly impact OS or risk of death in patients with stage I-II NSCLC. Patients harboring a *KRAS* G12C mutation did not have a worse OS. Taken together, these findings show good representativeness of this well-defined patient cohort.

Our study included only patients with stage I-II disease due to the focus on primary tumor size and to limit the prognostic impact of local invasion and regional lymph node involvement. Only 5.8% of the patients had N1 disease that could affect the prognosis. The major portion of the patients had tumor resection and more than 90% of tumors were adenocarcinoma. During this period, patients diagnosed with squamous cell carcinoma were molecularly assessed to a lesser extent, thus our study is more representative of adenocarcinoma. Even though most tumors were classified according the TNM staging guidelines 7^th^ edition, changes included in the 8^th^ edition, mainly covering substages that were not analyzed in this study, do not alter our findings [4].

When combining tumor size and *KRAS* mutational status there was no increased risk of death. No significant differences were observed when comparing OS for all stage I-II patients stratified by *KRAS* mutational status. However, the mean OS was 11 months shorter for *KRAS*^MUT^ patients. The same trend was observed when looking at resected patients with a 13-month shorter mean survival for *KRAS*^MUT^ patients. In this case, our results could differ from former studies due to earlier studies not being corrected for tumor size [15, 16]. Our cohort could be used as a historical reference if, in the future, sotorasib would be approved for adjuvant treatment for patients harboring *KRAS* G12C mutation.

Outcome variables other than survival such as recurrence rates and progression-free survival were not examined here. In addition, there remain confounders that were not include in the analyses such as the effect of different treatment methods on survival. Further, we use the T descriptor of the TNM staging system for tumor size even though the descriptor also includes invasion status and or intrapulmonary metastasis. We found that larger tumor size measured from resection specimens, but not CT scans, is associated with a higher risk of death. However, one confounder here is that non-resected patients are included in the CT group but not in the PAD group, which biases towards worse prognosis.

Going forward, much remains to be explored on the role of *KRAS* mutation in early NSCLC. In the age of precision medicine, our study contributes towards the detailed level clinical data that is required for future pooled analysis of prognosis assessments that can help guide clinical decisions.

In conclusion, we confirm the importance of primary tumor size and stage as a prognostic factor for survival in stage I-II NSCLC. *KRAS* mutations were not found to impact OS and no difference in the risk of death was observed when combining *KRAS* mutations and primary tumor size.

## Abbreviations

CT: Computed Tomography
ECOG: Eastern Cooperative Oncology Group
HR: Hazard Ratio
NSCLC: Non-Small Cell Lung Cancer
NGS: Next Generation Sequencing
PS: Performance Status
OS: Overall Survival

## Declarations

## Acknowledgments

We thank Sayin lab members and Nesrin Vurgun, Scientific editor at the Institute of Clinical Sciences, University of Gothenburg for a critical review of the manuscript. In addition, we thank members of the Swedish Lung Cancer Registry and the continuous reporting by Swedish healthcare employees.

## Authors’ contributions

Conceptualization, E.A.E., H.F., A.H. and V.I.S.; Data curation, E.A.E., A.M., H.F.; Formal analysis, E.A.E., A.M. and C.W.; Funding acquisition, E.A.E., C.W., H.F., A.H. and V.I.S.; Methodology, E.A.E.; Resources, H.F.; Supervision, A.H. and V.I.S.; Visualization, E.A.E., S.I.S., C.W. and V.I.S.; Writing—original draft, E.A.E., S.I.S and V.I.S.; Writing—review & editing, E.A.E., S.I.S, C.W., H.F., A.H. and V.I.S., Project coordination, V.I.S. All authors have read and agreed to the published version of the manuscript.

## Institutional Review Board Statement

Approval from the Swedish Ethical Review Authority (Dnr 2019-04771 and 2021-04987) was obtained prior to the commencement of the study. No informed consent was required due to all data presented in a de-identified form according to the Swedish Ethical Review Authority.

## Consent for publication

Not applicable. Patient consent statements were not required due to the retrospective nature of this study. No informed consent was required due to all data presented in a de-identified form according to the Swedish Ethical Review Authority.

## Data Availability Statement

The datasets used and/or analysed during the current study available from the corresponding author on reasonable request.

## Figure legends

**Supplemental Figure 1**

Kaplan-Meier estimates of overall survival for resected Stage I-II NSCLC patients stratified by KRAS mutational status. (**A**) No mutation in *KRAS* (wildtype, *KRAS*^WT^), with all *KRAS* mutations (*KRAS*^MUT^). (**B**) Only *KRAS*-*G12C* mutations (*KRAS*^MUT G12C^), *KRAS* mutations other than *G12C* (*KRAS*^MUT not G12C^).

**Supplemental Figure 2**

Kaplan-Meier estimates comparing overall survival for (**A**) all patients, (**B**) patients with no mutation in *KRAS* (wildtype, *KRAS*^WT^), and (**C**) with all *KRAS* mutations (*KRAS*^MUT^), stratified by Stages I and II.

**Supplemental Figure 3**

Kaplan-Meier estimates comparing overall survival for (**A**) all patients, (**B**) patients with no mutation in *KRAS* (wildtype, *KRAS*^WT^), and (**C**) with all *KRAS* mutations (*KRAS*^MUT^), stratified by TNM-stages T1, T2 and T3.

