## Supplementary material for "Combined assessment of *KRAS* mutational status and tumor size has no impact on prognosis in early-stage non-small cell lung cancer": Supp Figures 1-3

### Eklund et al. Supplemental Figure 1

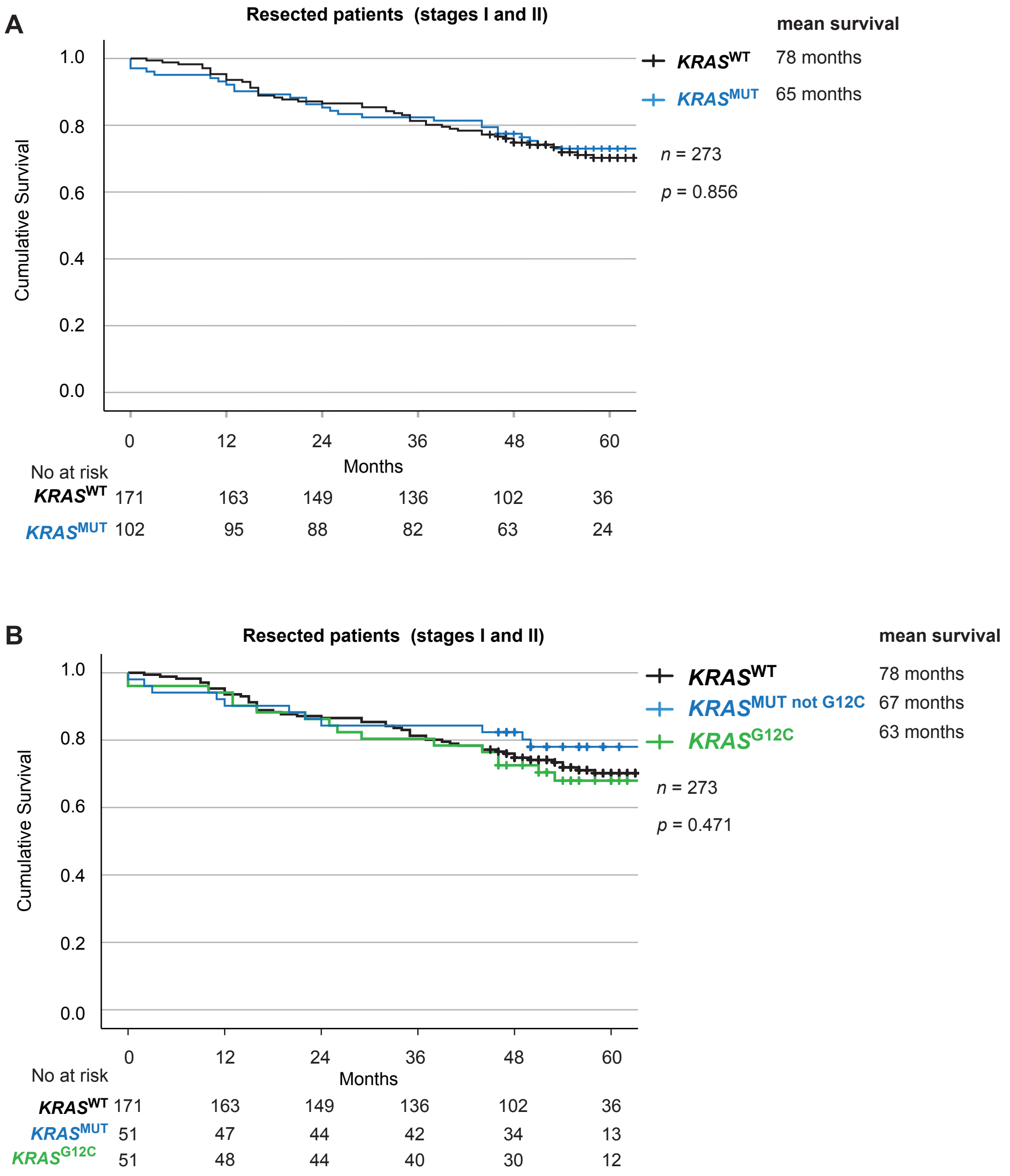

### Eklund et al. Supplemental Figure 2

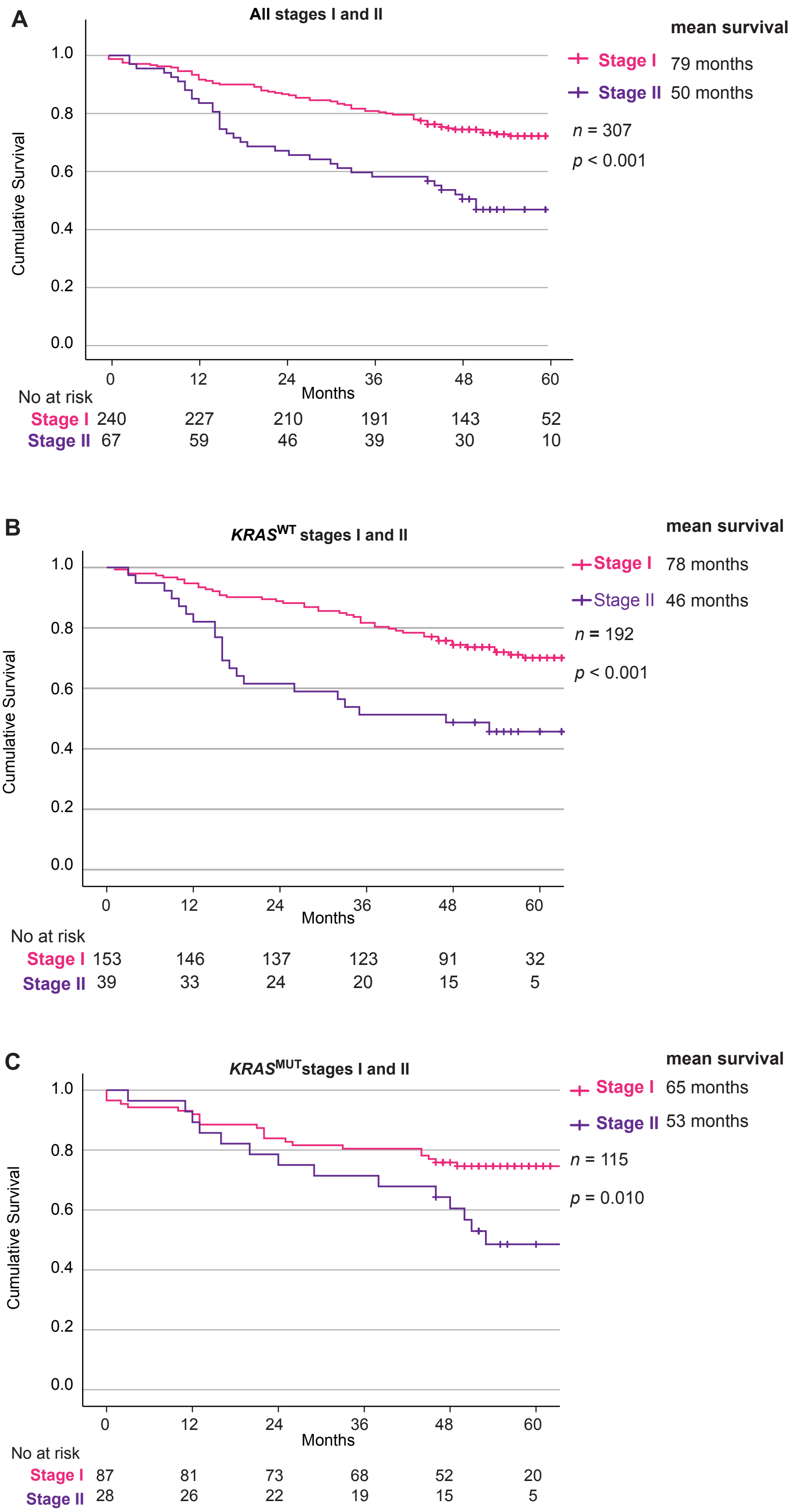

### Eklund et al. Supplemental Figure 3

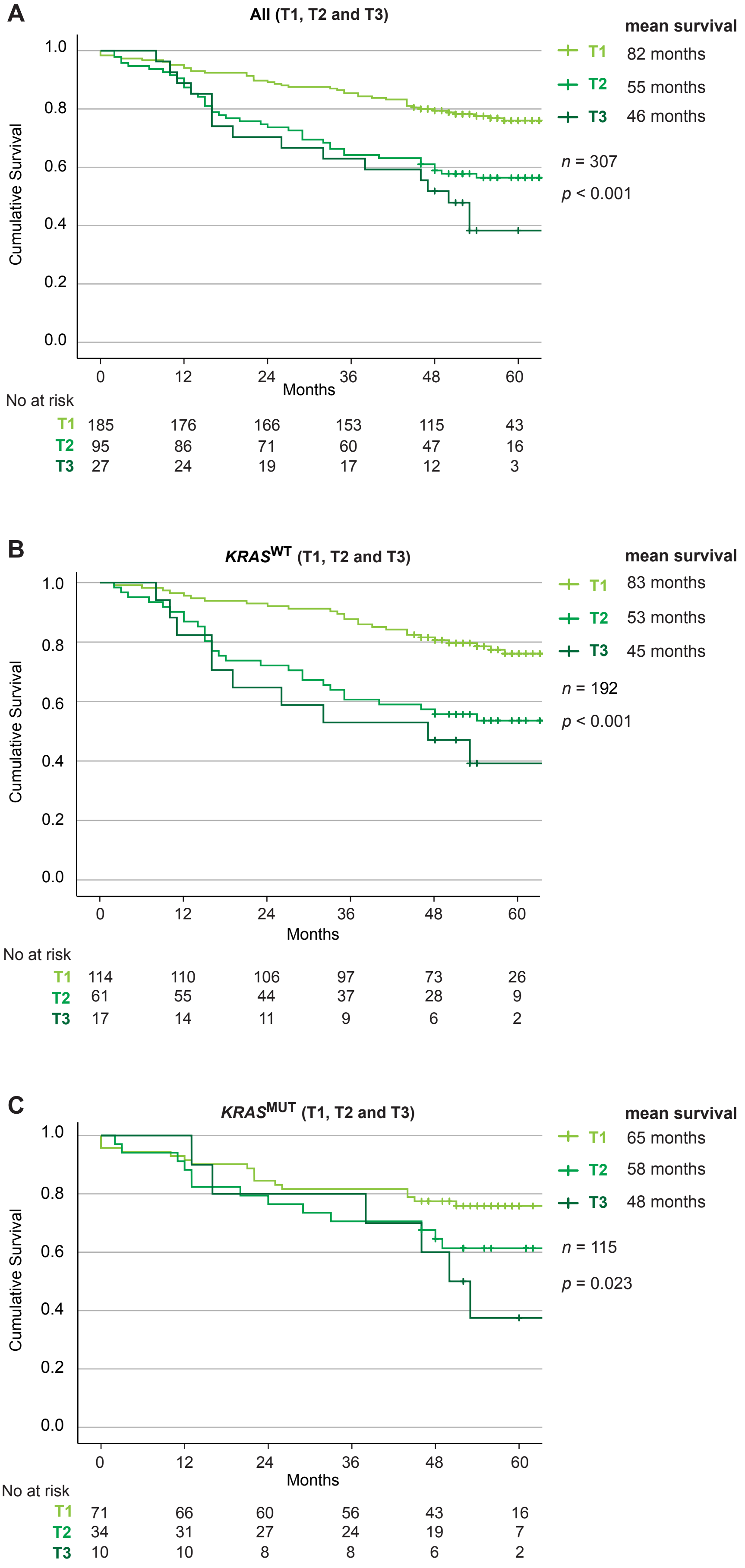
